# Quantifying the Effect Size of Exposure-Outcome Association Using *δ*-score: Application to Environmental Chemical Mixture studies

**DOI:** 10.1101/2022.03.02.22271732

**Authors:** Vishal Midya, Jiangang Liao, Chris Gennings, Elena Colicino, Susan L. Teitelbaum, Robert O. Wright, Damaskini Valvi

## Abstract

Epidemiologists often study the associations between a set of exposures and multiple biologically relevant outcomes. Frequently used scale and context dependent regression coefficients may not offer practically meaningful comparison and can further complicate the interpretation when these outcomes do not have similar units. Additionally, while scaling up a hypothesis driven study based on preliminary data, epidemiologists face a major uncertainty of how large the sample size should be. Frequently used p-value based sample size calculation emphasizes on precision and might lead to very large sample size for small or moderate effect sizes. This is not only costly but also might allow detection of irrelevant effects. Here we introduced a new concept “*δ*-score” by modifying Cohen’s *f* ^2^. *δ*-score is scale independent and under a new hypothesis testing framework quantifies the maximum Cohen’s *f* ^2^ with certain optimal properties. We also introduced “Sufficient sample size”, which is the sample size required to attain the *δ*-score. Finally, we demonstrated the utility of *δ*-score and sufficient sample size using data on adults from 2017–2018 US National Health and Nutrition Examination Survey for association between mixture of Per-and poly-fluoroalkyl substances and metals with serum high-density and low-density lipoprotein-cholesterols.

## 1 Introduction

Estimating effect size with considerable high precision is the currency of epidemiological research. Given a hypothesis of interest and a set of specific aims, preliminary data is collected. Collecting data and processing biological samples is costly and therefore this stage protects against waste of resources when the study does not progress as planned. Next depending on the obtained effect estimate and resource constraints, a larger study is planned. For example, consider a scenario where an epidemiologist wants to study the association between Perfluoroalkyl substances (PFAS) and alanine aminotransferase (ALT), a liver enzyme and cytokeratin-18 (CK-18), a marker of liver-cell death, in school age children. PFAS belong to a diverse class of environmental pollutants with “emerging concern” that interfere with multiple metabolic and hormonal systems in human (Futran Fuhrman et al. (2015)). ALT and CK-18 may or may not be measured in similar units and quantify different aspects of liver injury. Although, animal studies have shown biologically plausible cause-effect relationship between PFAS exposure and increase in ALT/CK-18 levels, their associations in humans are not well studied (Cano et al. (2021)).

Now assume, the regression estimates for both the associations be two unit increase per one unit increase in PFAS. If the units of ALT and CK-18 are different, the comparison between these estimates become difficult. Even a conversion to scale free outcomes makes the interpretation non-intuitive. Further, if ALT and CK-18 are on the same scale, two unit increase in ALT has a very different clinical and practical implication than two unit increase in CK-18, with one having higher potential for public health intervention than the other. Moreover, assume that at the current sample size, none of these associations are statistically significant. Based on their hypothesis, the epidemiologist decides to scale up their studies and apply for a grant based on their preliminary data. A p-value based sample size calculation yields large and comparable sample sizes and corresponding effect sizes become statistically significant. This situation leads to some quandaries, firstly, the increase in precision due to increase in sample size, may not reflect practically meaningful effect size and rather guarantees that any irrelevant and tiny effect sizes are detectable (Ioannidis et al. (2014), Wasserstein and Lazar (2016)). Secondly, for practically meaningful and statistically significant effect size, the high precision induced by large sample size may not be needed, as long as the effect size do not change considerably as sample size increases. Measuring PFAS/ALT/CK-18 in child serum is a time consuming and costly process, therefore even at a small/moderate sample size if the effect estimate allows for a contextual, biological and/or clinical implication, there may not be any further need to increase the sample size without a strong justification for higher precision.

A long established index to report strength of explanatory association in a more fundamental way is Cohen’s *f* ^2^ (Cohen (1988)), which evaluates the impact of additional variables over the baseline covariates. Through the past three decades, Cohen’s *f* ^2^ continues to be extensively used in behavioral, psychological and social sciences, due to its immense practical utility and ease of interpretation (Schäfer and Schwarz (2019)). In this paper, we propose *δ*-score by modifying Cohen’s *f* ^2^ to evaluate strength of explanatory association in a more fundamental and scale-independent way. Similar to Cohen’s *f* ^2^, *δ*-score moves the contextual reference to baseline covariates and evaluates the effect size contributed solely by a set of exposures or exposure-mixtures on top of those baseline covariates. Further, under a special hypothesis testing framework, we show that *δ*-score quantifies the maximum Cohen’s *f* ^2^ and admits some useful optimal properties. The idea was naturally extended to a new concept “Sufficient sample size” which is an estimate of the sample size required to attain the *δ*-score. Through illustrative examples and application in 2017–2018 US National Health and Nutrition Examination Survey (NHANES) data, we quantify *δ*-scores and Sufficient sample sizes for the association between mixture of Per-and poly-fluoroalkyl substances and metals on lipoprotein-cholesterols and demonstrate that Sufficient sample sizes are usually smaller than the p-value based sample size estimates.

## 2 Methods

Consider a common problem of testing if a set of exposures in a regression model is associated with the outcome after adjusting for covariates and/or confounders. For example, consider a linear model, ***y***=***X***_**0**_***b***_**0**_ + ***X***_**1**_***b***_**1**_ + ***ε***, and we are interested to know the strength of the association of ***X***_**1**_ after adjusting for ***X***_**0**_ and formulate the hypothesis, *H*_0_ : Effect size of ***X***_**1**_ = 0 vs *H*_1_ : Effect size of ***X***_**1**_ = *δ*, where *δ* is a pre-defined meaningful quantity and *δ* > 0. In the sections below, we first briefly discuss Cohen’s *f* ^2^ in linear regression. Next we move on to formulate an error calibrated hypothesis testing framework. Lastly, we introduce the idea of *δ*-score and Sufficient sample size under that framework.

### 2.1 Cohen’s *f* ^2^ in Linear Regression

Consider the linear regression model noted above and assume *ϵ* ∼ N (**0**, *σ*^2^***I***_***n***_), where ***I***_***n***_ is an identity matrix of dimension *n* × *n*. Let *γ*_*n*_ be the non-centrality parameter, then *γ*_*n*_ equals 0 when ***y*** is generated under *H*_0_. When ***y*** is generated under the alternative, *γ*_*n*_ has the form of 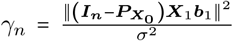, where 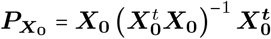 is the projection matrix on to the linear space spanned by the column vectors of ***X***_**0**_ (Wilks (1938), Brown et al. (1999)) (section S.1 of supplementary material). For the common regression design in which the predictor vector of each subject is drawn from a common population, *γ*_*n*_ grows linearly on *n*. Note that *γ*_*n*_ does not depend on *y* but depends on the design matrix *X* and underlying parameter *b*_1_ and *σ*^2^. A long established index of quantifying additional impact in linear regression is Cohen’s *f* ^2^,

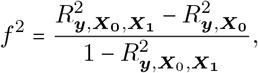

Where 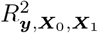 and 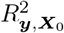 are the squared multiple correlation for ***X***_**0**_, ***X***_**1**_ under *H*_1_ and ***X***_**0**_ under *H*_0_ respectively. The *f* ^2^ quantifies the proportion of association with ***y*** accounted by ***X***_**1**_ on top of the association accounted by ***X***_**0**_, a concept that most researcher can relate to intuitively (Selya et al. (2012)). In linear regression, Cohen’s *f* ^2^ and non-centrality parameter *γ*_*n*_ can be connected through the following lemma.

#### Lemma 1

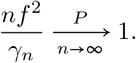

See the proof in section S.2 of supplementary material. Further discussion on Cohen’s *f* ^2^ in generalized linear models is presented in section S.3 of supplementary material.

### 2.2 Formulation of error calibrated cutoff in a new hypothesis testing framework

Following the hypothesis in (1) and for a meaningful value of *δ* > 0, we specify our main hypothesis as below

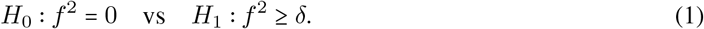

Let the test statistic be *S* (***y***) = *p*_1_*F* (***y***) for a linear regression and *S*(***y***) = Λ (***y***) for other generalized linear models. T be a type 1 and type 2 error calibrated cutoff which depends on sample size *n* and unknown parameters *p*_1_ and effect size *δ*. Then given cutoff T, one can define a testing procedure by its type 1 and type 2 errors as below:

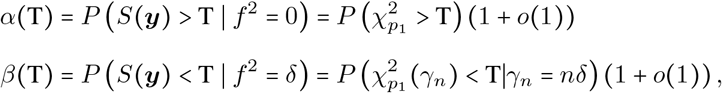

Where 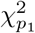 denotes central chi-squared random variable with *p*_1_ degrees of freedom and 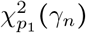 denotes non-central chi-squared random variable with *p*_1_ degrees of freedom and *γ*_*n*_ as the non-centrality parameter. Our central idea is to choose T so that type 1 error *α* (T) and the type 2 error *β* (T) satisfy the relationship, *α*(T) = *θβ*(T), with 0<*θ*≤1 and *θ* is pre-specified. Using the chi-square approximation to *S* (***y***), we can solve for the calibrated cutoff T by equation

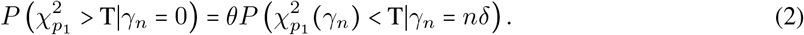

When T is fixed, the left size of equation (2) remains constant as *n*→ ∞ while the right side diminishes to 0 rapidly under non-centrality parameter *nδ*. Therefore, equation (2) implies T→ ∞ as *n*→ ∞. In the Theorem stated below we elaborate more on T. The results in theorem 1 depend on the normality approximation of the non-central chi-square distribution, i.e. for large *n*, equation (2) was rewritten as,

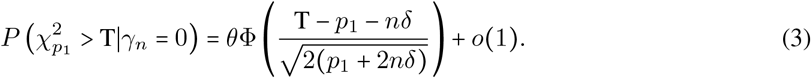

For ease of interpretation and theoretical derivations, we consider *θ*= 1 in the following sections, i.e. the case when both the type 1 and type 2 errors decay at the same rate. The cases with *θ*≠ 1 can be developed similarly.

#### Theorem 1.

*Consider the hypothesis of interest to be H*_0_ : *f* ^2^ = 0 *vs. H*_1_ : *f* ^2^ ≥ *δ, where f* ^2^ *denotes Cohen’s f* ^2^. *Assume data* ***y*** *is generated under the alternative with f* ^2^ = *δ. Then following the constraint α*(*T*) = *β*(*T*) *as in (2) and for large n, the error calibrated cutoff T has the following expression*,

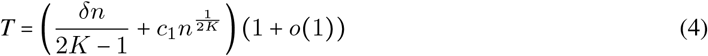

*Further, the type 1 error or the type 2 error rates can be expressed as*,

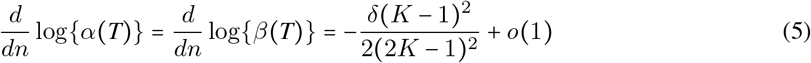

*where*, 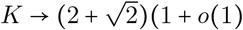 *and c*_1_ *is a constant of integration*.

The proof is presented in section S.4 of supplementary material. Theorem 1 sheds light on the structure of the cutoff T and the rates of corresponding type 1 or type 2 errors when the sample size *n* is large. Since both errors go to 0 as *n* → ∞, this procedure for testing of hypothesis is consistent while keeping error rates equal. It should be noted that both the errors decay at an exponential rate and therefore deems useful even at moderate sample sizes. In order to convince the accuracy of Theorem 1, we presented the type 1 and type 2 error rates as well as the rate of change of T with respect to *n* using the results from theorem 1 and corresponding numerical results from equation (3). As seen from Table 1, irrespective of the Cohen’s *f* ^2^, as *n* increases, the rate of change of T, log (type 1) and log (type 2) converge to the corresponding theoretical rates specified in Theorem 1. We also conducted a Monte Carlo simulation to estimate the calibrated type 1 and type 2 errors for different values of *n, p*_1_ and *f* ^2^ (see section S.5 and Table 1 of supplementary material). Moreover, detailed discussion on the properties of the error calibrated cutoff T and the type 2 error is presented in section S.6 of supplementary material.

**Table 1:** Rates of cutoff T, log (type 1) or log (type 2) with respect to sample size *n*, based on Theorem 1 and equation (3)

|  |  |  | Numerical approximation using Equation (3) |  | Using Theorem (1) |  |
| --- | --- | --- | --- | --- | --- | --- |
| $p_1$ | $f^2$ | $n$ | $\frac{d}{dn} T$ | $\frac{d}{dn} \log(\alpha)$ | $\frac{d}{dn} T$ | $\frac{d}{dn} \log(\alpha)$ |
| 1 | 2.5% | 250 | 0.0042895 | -0.0031 | 0.0042893 | -0.0021 |
| 1 | 10% | 250 | 0.0171573 | -0.0100 | 0.0171573 | -0.0086 |
| 5 | 2.5% | 250 | 0.0043764 | -0.0025 | 0.0042893 | -0.0021 |
| 5 | 10% | 250 | 0.0172491 | -0.0090 | 0.0171573 | -0.0086 |
| 1 | 2.5% | 500 | 0.0042896 | -0.0028 | 0.0042893 | -0.0021 |
| 1 | 10% | 500 | 0.0171573 | -0.0094 | 0.0171573 | -0.0086 |
| 5 | 2.5% | 500 | 0.0043764 | -0.0024 | 0.0042893 | -0.0021 |
| 5 | 10% | 500 | 0.0172491 | -0.0087 | 0.0171573 | -0.0086 |

### 2.3 Notion of *δ*-score

We can borrow the common convention for *f* ^2^ (Cohen (1988)) and call *f* ^2^≥0.02, *f* ^2^≥0.15 and *f* ^2^≥0.35 as representing small, moderate and large effect size respectively. This can serve as the guidance in understating the effect size obtained from the data. Further, given the data, one can utilize this hypothesis by sequentially choosing and testing increasing values of *δ* as long as the null gets rejected and finally stopping when the null can no longer be rejected. Finally, this brings us to the question that given any data whether there exists any maximum *δ* such that the null will always be rejected. Let the likelihood ratio test statistic be Λ(***y***). We reject the null if and only if T (*δ*) ≤ Λ (***y***). Hence, T (*δ*) attains a maximum at the upper bound Λ (***y***). Denote this *δ* at which T (*δ*) attains the maximum as *δ*^∗^ and consider the reformulated hypothesis (1) as below, with *δ*^∗^ as the final choice of *δ*,

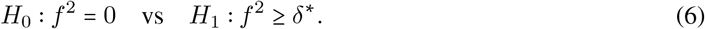

We note some interesting properties of *δ*^∗^ through the following corollary,

#### Corollary 1

*Under the hypothesis in (6), let δ*^∗^ *be the unique solution to the equation T*(*δ*^∗^)= Λ (***y***). *Then δ*^∗^ *admits the following properties:*

- *δ*^∗^ *is the maximum value of Cohen’s f* ^2^ *such that the null is still rejected*.
- *For any h* ≥ −*δ*^∗^, *the asymptotic type 1 error*, 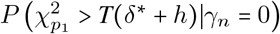 *is a monotonically decreas-ing function of h, whereas the asymptotic type 2 error*,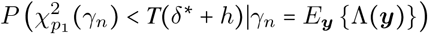 *is a monotonically increasing function of h*.

See the proof in Section S.8 of supplementary material. Further, for large *n, δ*^∗^ undertakes asymptotic convergence (Lemma 3.1 of Vuong (1989)) and we define “*δ*-score” as noted below:

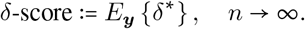

Under the hypothesis testing framework in (1), *δ*-score captures the asymptotic and maximum Cohen’s *f* ^2^ contributed solely by the larger exposure model on top of baseline covariate only model. Unlike usual Null Hypothesis Significance Testing based sequential testing (Schönbrodt et al. (2017)), this framework does not inflate the type 1 error and circumvents the issue through its use of the error calibrated cutoff and keeps the error rates in balance. Instead of simply estimating Cohen’s *f* ^2^, this procedure introduces hypothesis testing for an error balanced decision making.

### 2.4 Notion of Sufficient sample size

*δ*-score can be estimated by bootstrapping a large size *N* (say *N*= 5000 or 10000) with replacement from the original sample of size *n*, with *n*< *N*. Moreover, because of its convergence, one can find a much smaller bootstrapped size and corresponding estimated *δ*-score such that it will be in a “practically close neighbourhood” of the converged *δ*-score based on very large bootstrap size. We define that smaller bootstrapped size as Sufficient sample size.

Consider the equivalence tests for the ratio of two means with prespecified equivalence bounds (Schuirmann (1987) and Phillips (1990)). Let *δ*^*s*^ and *δ*^opt^ be the underlying random variables for two separate *δ*-scores to be estimated under sample sizes *N* and *n*_*s*_ respectively. To formulate the test of non-equivalence between these two estimated *δ*-scores, consider the hypothesis below,

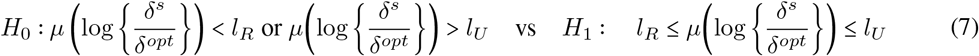

where, *l*_*R*_ and *l*_*U*_ are the lower and upper equivalence bounds with *l*_*R*_ < 0 and *l*_*U*_ > 0. The null hypothesis will be rejected to favour the alternative if a two-sided 100 (1−2*α*) % CI is completely included within *l*_*R*_ and *l*_*U*_. We will assume *l*_*R*_ = log(0.8) and *l*_*U*_ = log(1.25) following typical practice (Phillips (2009)) but less stricter values can be chosen for practical purposes. 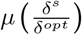 and 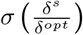 are approximated by using Taylor series expansions (detailed in Section S.2 of supplementary material). The mean and variance after logarithmic transformation are found using direct application of delta theorem on 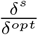. Finally, we declare alternative hypothesis if the 2*α* level CI on 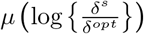 is within the equivalence limits, i.e.,

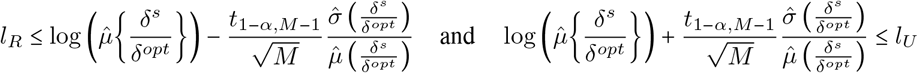

where, *t*_1−*α,M*−1_ is the 100 (1−*α*)%^th^ quantile in a standard t-distribution. As long as the hypothesis of non-equivalence in (7) is rejected in favour of the alternative, *n*_*s*_ can be regarded as a “sufficient sample size” at equivalence bounds of 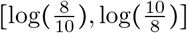 with a corresponding *δ*-score of 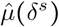.

## 3 Illustration with a simulated example

Consider a normally distributed outcome and one single exposure with five baseline covariates with sample size of 300. Further assume, the *R*^2^ for the baseline covariate only model is 20%, and the true and unknown *δ*-score due to the exposure is 5.8%. Therefore, the *R*^2^ for the larger model with a single exposure and five covariates is 20.8% (the mean correlation between the covariates is set at 0.3 and the error variance is assumed to be 5). See Section S.7 of the supplementary material for the data generating process.

Assume a researcher collected this data and intends to find the association between the outcome and the exposure after controlling for the five baseline covariates. As a first step, *δ*-score is estimated by bootstrapping a size *N* = 5000 from original sample of *n* = 300. The estimated Impacts score is 6.1% (which is very close to the true impact of 5.8%). Similarly, *δ*-scores are estimated at bootstrapped sizes *N*=200, 300, 400, 500, 600 and 2500 to illustrate the gradual convergence as the bootstrap size increases (Fig. 1-A). Further note that, even when precision increases with bootstrap size, the mean of regression coefficients remain stable (Fig. 1-B) while the p-values keep getting smaller (Fig. 1-D).

**Figure 1:**
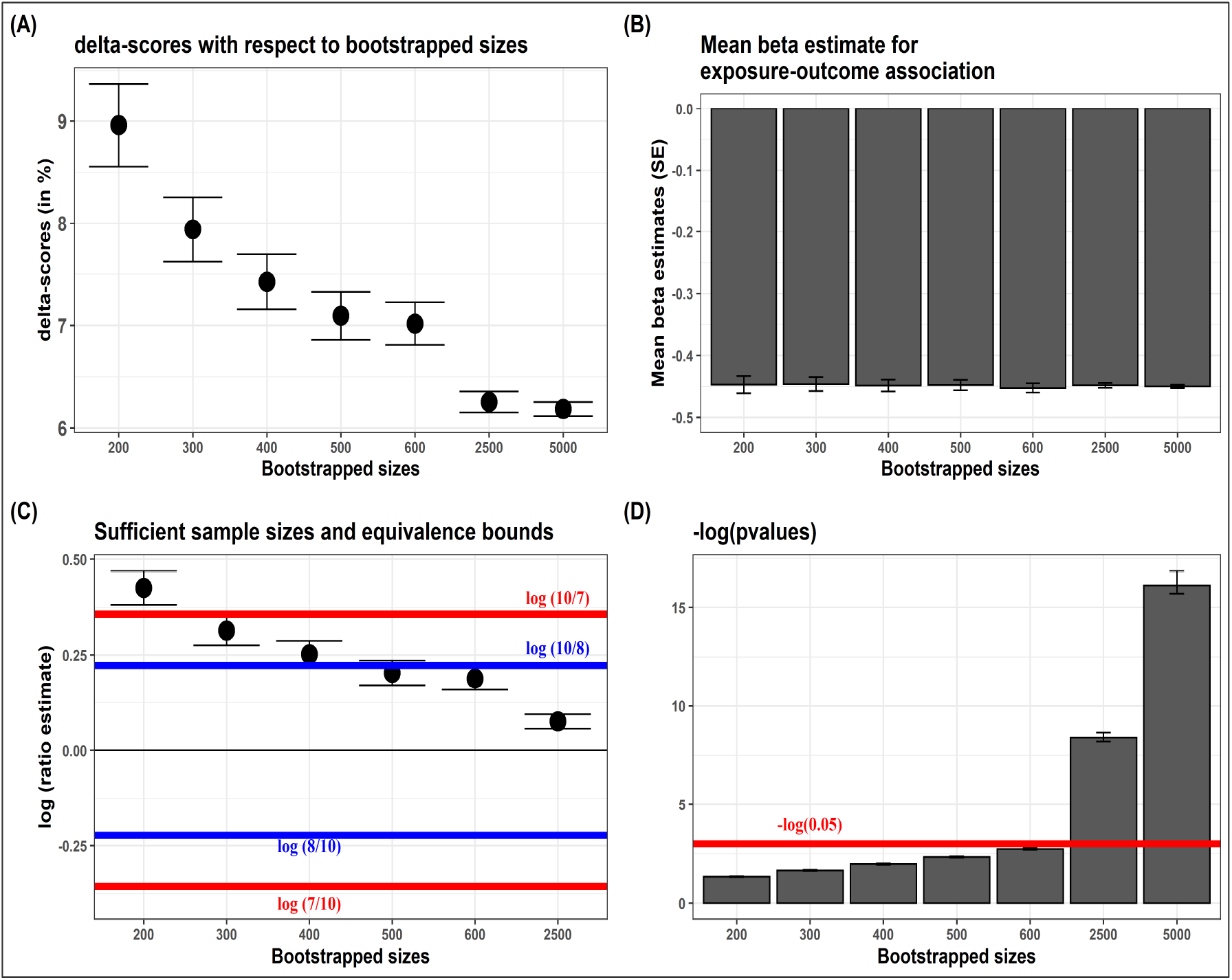
Results from simulated example. (A) Illustration of *δ*-scores for different bootstrapped sizes and its eventual convergence, (B) Mean *β* estimates and Standard errors of the exposure-outcome association, (C) Sufficient sample sizes with respect to choices of equivalence bounds (D) Negative log (base = e) p-values as bootstrapped sizes increase

For the original sample size of *n*=300, the corresponding p-value of the regression estimate of the exposure, is not significant. The researcher therefore might want scale up the study to collect more data and increase the original sample size based on statistical power calculation and sample size determination - which estimates that a total sample size of around 1000 is required assuming 80% power and type 1 error fixed at 5%. Sufficient sample size estimation using *δ*-score strikes a balance between precision and utility. We estimated 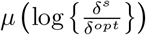 based on 2000 iterations and used the hypothesis of non-equivalence in (7) to compare the *δ*-scores at *N* = 200, 300, 400, 500, 600 and 2500 with respect to the estimated *δ*-score at *N* = 5000 (Fig. 1-C). At *N* = 600, the 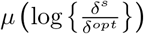 and its 95% CI lies within the bounds of *l*_*R*_ =log (8/10) and *l*_*U*_ = log (10/8), whereas at *N* =500, 400 and 300, it breaches the upper bound of *l*_*U*_ =log (10/8) but stays within the bounds of *l*_*R*_ =log (7/10) and *l*_*U*_ =log (10/7). Accordingly, the researcher can choose a sufficient sample size of *N* =600 or *N* =300 at equivalence bounds of [log (8/10), log (10/8)] or [log (7/10), log (10/7)]respectively with corresponding *δ*-scores 7% and 7.9%. These *δ*-scores are within a close neighbourhood of the converged *δ*-scores of 6% (based on the bootstrapped size of *N* = 5000).

## 4 Application in exposure-mixture association of PFAS and metals with serum lipids among US adults

PFAS are exclusively man-made EDCs and environmentally persistent chemicals which are used to manufacture a wide variety of consumer and industrial products, non-stick, stain and water resistant coatings, fire suppression foams, and cleaning products (Liu et al. (2018) and Jain and Ducatman (2018)). Both PFAS and metals have been associated with increase in cardiovascular disease (CVD) or death using many crosssectional and longitudinal observational studies and experimental animal models (Meneguzzi et al. (2021)). Hypercholesterolemia is one of the significant risk factors for CVD and it is characterized by the presence of high levels of cholesterol in the blood. High serum low-density lipoprotein (LDL), total serum cholesterol levels, and low levels of high-density lipoprotein (HDL) in the blood are one of the incriminating factors for the pathogenesis of this disorder (Buhari et al. (2020)). Using the theory discussed in the sections above, we aim to quantify the *δ*-scores of PFAS and metal mixtures on serum lipoprotein-cholesterols and estimate sufficient sample sizes.

### 4.1 Study Population

We have used a cross-sectional data from the 2017–2018 US NHANES (CDC and NCHS (2018)). The present study has data on 683 adults. Data on baseline covariates (age (in years), gender, ethnicity, body mass index (bmi) (in *kg*/*m*^2^), smoking status, ratio to family income to poverty) were downloaded and matched by IDs of the NHANES participants. See Table 2 for details on characteristics of the study population. To adjust for oversampling of non-Hispanic black, non-Hispanic Asian, and Hispanic in NHANES 2017-2018, a weight variable was added in the regression models. List of individual PFAS, metals and their lower limit of detection can be found in section S.9 in the supplementary material.

**Table 2:** Study characteristics of the population under investigation - data from National Health and Nutrition Examination Survey 2017–2018

|  | Total | Male | Female | % observations ≥ LLOD |
| --- | --- | --- | --- | --- |
| Sample size (n) | 683 | 339 | 344 |  |
| Baseline Covariates |  |  |  |  |
| Age (years) | 49.51 (18.77) | 50.38 (18.81) | 48.65 (18.73) |  |
| Ethnicity |  |  |  |  |
| Mexican American | 88 | 43 (49%) | 45 (51%) |  |
| Other Hispanic | 58 | 23 (40%) | 35 (60%) |  |
| Non-Hispanic White | 260 | 135 (52%) | 125 (48%) |  |
| Non-Hispanic Black | 155 | 79 (51%) | 76 (49%) |  |
| Other Race - Including Multi-Racial | 122 | 59 (48%) | 63 (52%) |  |
| Body mass index ($kg/m^2$) | 29.59 (7.90) | 28.67 (6.36) | 30.49 (9.09) | |
| Smoking Status |  |  |  |  |
| Never | 402 | 170 (42%) | 232 (58%) |  |
| Smoked at least 100 cigarettes in life but don't smoke now | 163 | 100 (61%) | 63 (39%) |  |
| Smoked at least 100 cigarettes in life and smoke now | 118 | 69 (58%) | 49 (42%) |  |
| Ratio of family income to poverty | 2.56 (1.61) | 2.64 (1.63) | 2.48 (1.59) |  |
| Outcomes |  |  |  |  |
| HDL-C (mg/dL) | 53.91 (15.53) | 49.19 (13.10) | 58.56 (16.33) |  |
| LDL-C (mg/dL) | 109.35 (37.11) | 108.99 (35.35) | 109.71 (38.83) |  |
| PFAS exposures (Unadjusted geometric means with 95% confidence intervals) |  |  |  |  |
| PFDeA (ng/mL) | 0.20 (0.19, 0.21) | 0.21 (0.19, 0.22) | 0.20 (0.18, 0.22) | 68.73 % |
| PFHxS (ng/mL) | 1.10 (1.03, 1.17) | 1.49 (1.38, 1.61) | 0.81 (0.74, 0.89) | 99.12% |
| Me-PFOSA-AcOH (ng/mL) | 0.13 (0.12, 0.14) | 0.14 (0.13, 0.15) | 0.12 (0.11, 0.13) | 38.64% |
| PFNA (ng/mL) | 0.42 (0.39, 0.44) | 0.46 (0.42, 0.5) | 0.38 (0.34, 0.42) | 91.74% |
| PFUA (ng/mL) | 0.14 (0.13, 0.15) | 0.14 (0.13, 0.15) | 0.14 (0.13, 0.15) | 41.59% |
| n-PFOA (ng/mL) | 1.28 (1.22, 1.35) | 1.52 (1.42, 1.64) | 1.08 (1, 1.17) | 99.41% |
| n-PFOS (ng/mL) | 3.26 (3.04, 3.5) | 4.11 (3.74, 4.51) | 2.59 (2.35, 2.86) | 99.41% |
| Sm-PFOS (ng/mL) | 1.28 (1.19, 1.37) | 1.73 (1.58, 1.89) | 0.95 (0.86, 1.04) | 98.82 % |
| Lead, Cadmium, Total Mercury, Selenium, & Manganese exposures (Unadjusted geometric means with 95% confidence intervals) |  |  |  |  |
| Cd ($\mu g/L$) | 0.32 (0.3, 0.34) | 0.29 (0.27, 0.32) | 0.35 (0.32, 0.38) | 91.36% |
| Pb ($\mu g/dL$) | 0.91 (0.86, 0.96) | 1.09 (1, 1.18) | 0.76 (0.7, 0.82) | 100% |
| Mn ($\mu g/L$) | 9.45 (9.21, 9.7) | 8.91 (8.62, 9.22) | 10.01 (9.64, 10.41) | 100% |
| THg ($\mu g/L$) | 0.78 (0.72, 0.84) | 0.81 (0.73, 0.9) | 0.75 (0.67, 0.83) | 84.77% |
| Se ($\mu g/L$) | 188.62 (186.75, 190.52) | 189.28 (186.57, 192.04) | 187.97 (185.38, 190.6) | 100% |

Data presented as mean(SD) or $n(\%)$ ; LLOD: lower limit of detection (in ng/mL); LDL-C: low-density lipoprotein-cholesterol (mg/dL) ; HDL-C: high-density lipoprotein-cholesterol (mg/dL); PFDeA: Perfluorodecanoic acid; PFHxS: Perfluorohexane sulfonic acid; Me-PFOSA-AcOH: 2-(N-methylperfluorooctanesulfonamido)acetic acid; PFNA: Perfluorononanoic acid; PFUA: Perfluoroundecanoic acid; PFDoA: Perfluorododecanoic acid; n-PFOA: n-perfluorooctanoic acid; Sb-PFOA: Branch perfluorooctanoic acid isomers; n-PFOS: n-perfluorooctane sulfonic acid; Sm-PFOS: Perfluoromethylheptane sulfonic acid isomers; Pb: Lead; Cd: Cadmium; THg: Total Mercury; Se: Selenium; Mn: Manganese

### 4.2 Methods

We will use Weighted Quantile Sum regression (Carrico et al. (2015)) but other exposure-mixture models such as Bayesian weighted quantile sum regression (Colicino et al. (2020)) and Bayesian kernel machine regression (Bobb et al. (2014)) can also be utilized, as long as the likelihood ratio test statistic can be estimated. All the PFAS and metals were converted to decile. As an additional analysis, both the serum cholesterols were also dichotomized using their 90^*th*^ percentile, to demonstrate the effectiveness of *δ*-scores on binary outcomes. *δ*-scores were estimated using bootstrapped sizes of 5000 from the original sample of size 683 and the process was iterated 100 times.

### 4.3 Results

For metals and PFAS, the *δ*-scores of continuous HDL-C were 9.6% [95% CI: (9.1%, 10.0%)] and 10.7% [95% CI: (10.2%, 11.1%)] respectively, whereas for continuous LDL-C, those were 14.7% [95% CI: (14.2%, 15.2%)] and 16.2% [95% CI: (15.6%, 16.7%)] respectively. Both the mixtures have relatively higher *δ*-scores on LDL-C than HDL-C. Further, for both the cholesterols, metal-mixture has slightly higher *δ*-score than the PFAS-mixture (Fig.2 - A and B). PFAS and Metal mixtures have higher *δ*-scores for LDL-C than HDL-C. Further, after dichotomizing both the cholesterols at their 90^*th*^ percentile, the *δ*-scores for metal-mixture remained similar to the continuous cholesterol outcome (HDL-C: 9.8% [95% CI: (9.4%, 10.2%)] and LDL-C: 17.2% [95% CI: (16.6%, 17.8%)]), but slightly decreased for PFAS-mixture (HDL-C: 6.9% [95% CI: (6.5%, 7.2%)] and LDL-C: 11.5% [95% CI: (11.0%, 12.0%)]). The decrease might have been due to some loss of information while dichotomizing the outcome (Fig.2 - C and D).

**Figure 2:**
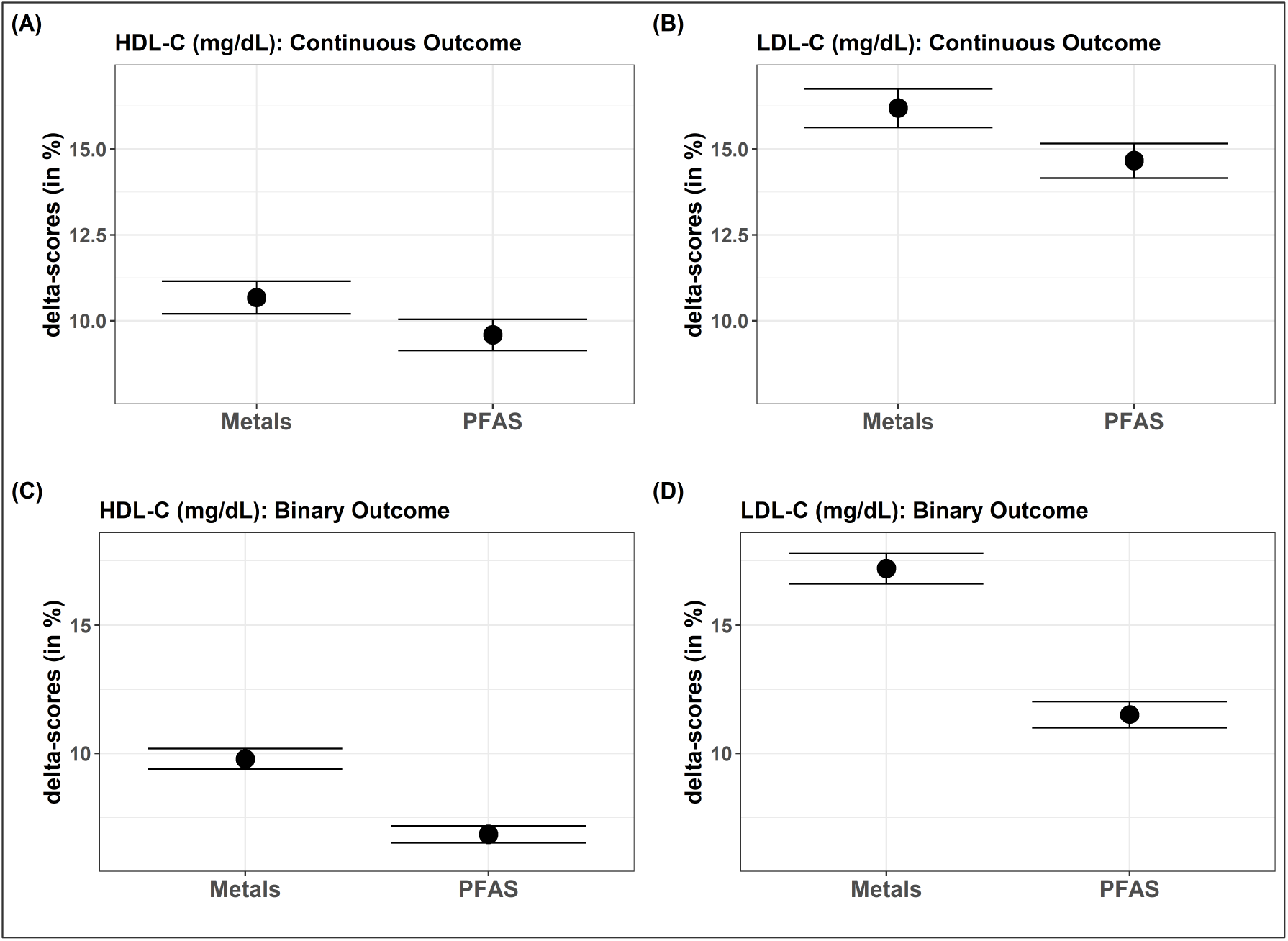
*δ*-scores of EDC exposure-mixture to quantify the variability in serum lipoprotein-cholesterols

Sufficient sample sizes were also estimated for this dataset at the equivalence bounds of 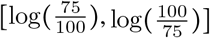. For both metal and PFAS-mixtures, the 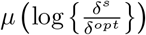 and their corresponding 95% CI for bootstrap size 683, lie well within the equivalence bounds. Further even at a decreased sample size of 483, the 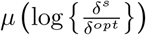 and their 95% CIs, still remain with the equivalence bounds. Therefore, *N* = 483, is a sufficient sample size at equivalence bounds 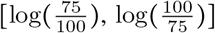 for both metal and PFAS-mixture (Fig.3). But further decrease in the bootstrap size, would not be sufficient, at this pre-fixed equivalence bounds. One can further modify the bootstrap size *N* = 483 to obtain a precise estimate of sufficient sample size.

**Figure 3:**
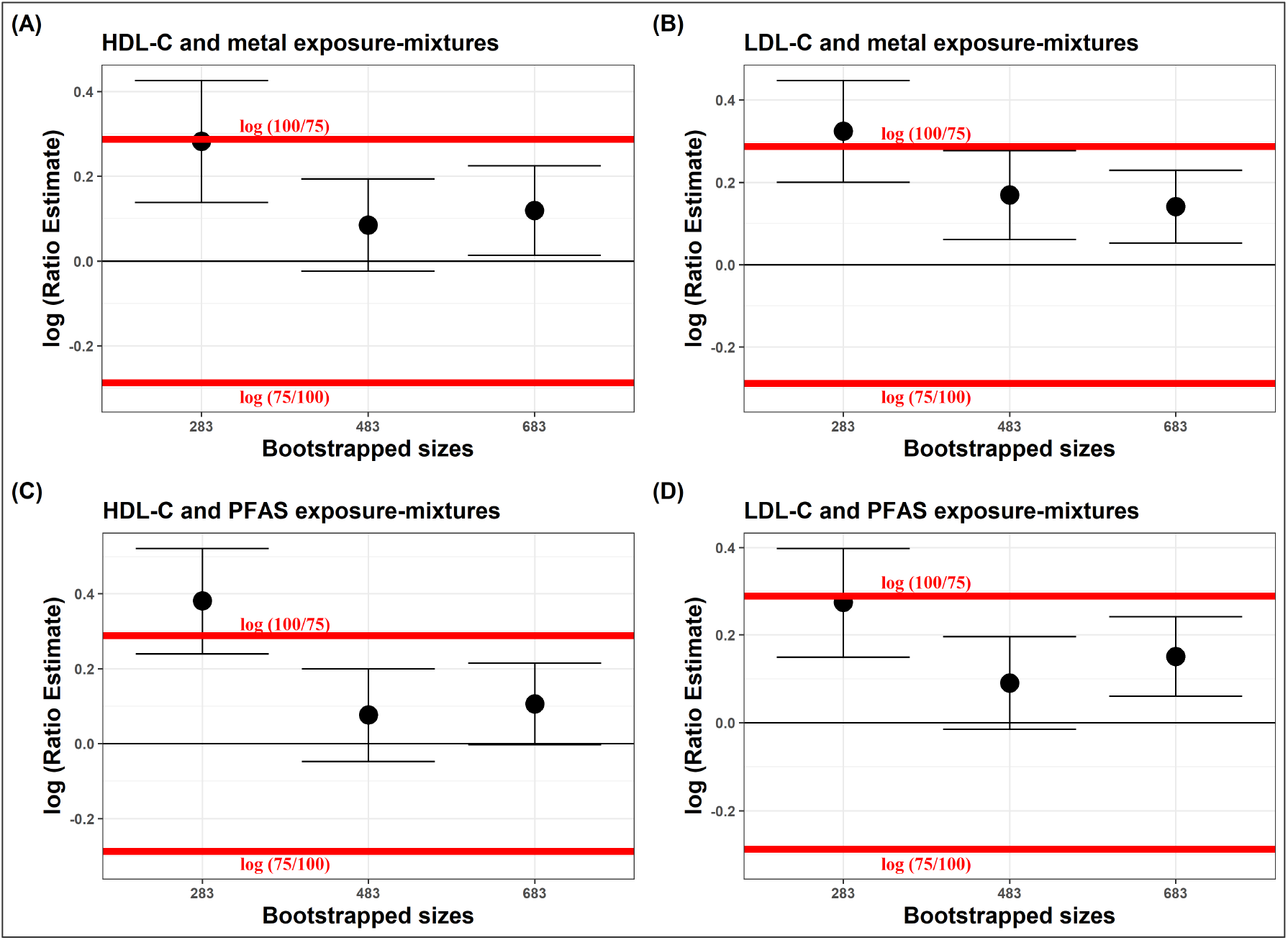
Sufficient sample sizes to estimate the *δ*-scores in serum lipoprotein-cholesterols explained solely by EDC exposure-mixture at equivalence bounds of 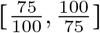

## 5 Concluding remarks

In this paper, we introduced the idea of *δ*-score for and Sufficient sample size for exposure-outcome association. *δ*-score is easily interpretable, scale independent and because of its connect to Cohen’s *f* ^2^, *δ*-score allows for direct comparisons between several outcomes measured on different scales or separate studies or in meta-analysis. Thereafter, *δ*-score can be potentially used to compare and choose between multiple outcomes with varying units and scales. Further, sample size determination based on preliminary data might utilize Sufficient sample size in designing more cost-efficient human studies.

This framework has its limitation, the bootstrapped estimation of *δ*-score assumes the original sample is well representative of the true target population. Any estimation of *δ*-score, therefore carries this implicit assumption. But such an assumption is at the core of many statistical analyses and a well designed study can ideally alleviate such issues or could be corrected to be well representative. In addition, this current theory is based on likelihood ratio test of nested models but future work can extend this framework to strictly nonnested or overlapping models. Progress can also be made to estimate *δ*-score on high dimensional setting which can be utilized in Metabolomics studies.

## Data Availability

All data produced are available online at US NHANES(2017 and 2018)

https://wwwn.cdc.gov/nchs/nhanes/continuousnhanes/default.aspx?BeginYear=2017

## 6 Supplementary Information

Supplementary material is provided in a separate file. All the R codes used in the article are available on GitHub.

## Notes

### Competing Interest Statement

The authors have declared no competing interest.

### Funding Statement

This study was funded by - National Institute of Environmental Health Science (NIEHS): P30ES023515

### Summary of Updates

1) The aims and presentation of the paper 2) Most technical details were moved to the supplement

