## Supplementary material for "Quantifying the Effect Size of Exposure-Outcome Association Using *δ*-score: Application to Environmental Chemical Mixture studies"

#### Contents

**1 Derivation of non-centrality parameter $\gamma_n$ in linear regression using Information Method 3**

**2 Proof of Lemma 1 4**

**3 Cohen's $f^2$ in Generalized Linear Models 5**

3.1 Connection between Cohen's $f^2$ and non-centrality parameter in Generalized Linear Models 6

**4 Proof of Theorem 1 7**

**7 Proof of Corollary 1 14**

**8 Data generating process for simulations 14**

9 PFAS and metal exposures and Serum lipoprotein-cholesterols outcomes 16

#### 1 Derivation of non-centrality parameter $\gamma_n$ in linear regression using Information Method

First note that the non-centrality parameter $\gamma_n$ is same for both the F-distribution and the Chi-squared distribution under linear regression. Brown et al. (1999) discussed the "Information Method" for computing $\gamma_n$. Assume that $y_1, \ldots, y_n$ be a realization of independent and identically distributed random variable $\boldsymbol{y} = \{y_1, \ldots, y_n\}$, with $\boldsymbol{y} \sim \mathcal{N}(\boldsymbol{X_0 b_0} + \boldsymbol{X_1 b_1}, \sigma^2 \boldsymbol{I_n})$ and $\sigma^2$ is known. Consider the hypothesis $H_0 : f^2 = 0 \quad \text{vs} \quad H_1 : f^2 = \delta$. Under the null, $\boldsymbol{b_1} = \boldsymbol{0}$ since $f^2 = 0$. The parameter $\boldsymbol{b_0}$ is not constrained by the hypothesis and therefore can be estimated by the unconstrained MLE. Let, $\ell(\boldsymbol{y}|\boldsymbol{X_0}, \boldsymbol{X_1})$ be the log-likelihood function for observations $\boldsymbol{y}$ and $\boldsymbol{i}$ be the expected information matrix which is partitioned with respect to $\boldsymbol{b_1}$ and $\boldsymbol{b_0}$ as,

$$\begin{pmatrix} \boldsymbol{i}_{b_1,b_1} & \boldsymbol{i}_{b_1,b_0} \\ \boldsymbol{i}_{b_0,b_1} & \boldsymbol{i}_{b_0,b_0} \end{pmatrix}.$$

Then the non-centrality parameter $\gamma_n$ can be then be expressed as below (For more details on this approximation, refer to Chapter 9 of Cox and Hinkley (1979)):

$$\gamma_n = \boldsymbol{b_1}^t \left( \boldsymbol{i}_{b_1,b_1} - \boldsymbol{i}_{b_1,b_0} \boldsymbol{i}_{b_0,b_0}^{-1} \boldsymbol{i}_{b_0,b_1} \right) \boldsymbol{b_1} \tag{1}$$

The expected information matrix $\boldsymbol{i}$ from the log-likelihood equation is:

$$\boldsymbol{i} = \begin{pmatrix} \boldsymbol{i}_{b_1,b_1} & \boldsymbol{i}_{b_1,b_0} \\ \boldsymbol{i}_{b_0,b_1} & \boldsymbol{i}_{b_0,b_0} \end{pmatrix} = \begin{pmatrix} \frac{\boldsymbol{X}_1^t \boldsymbol{X}_1}{\sigma^2} & \frac{\boldsymbol{X}_1^t \boldsymbol{X}_0}{\sigma^2} \\ \frac{\boldsymbol{X}_0^t \boldsymbol{X}_1}{\sigma^2} & \frac{\boldsymbol{X}_0^t \boldsymbol{X}_0}{\sigma^2} \end{pmatrix} \tag{2}$$

Combining 1 and 2, we obtain

$$\begin{aligned} \gamma_n &= \frac{1}{\sigma^2} \left[ (\boldsymbol{X_1 b_1})^t (\boldsymbol{X_1 b_1}) - (\boldsymbol{X_1 b_1})^t \left( \boldsymbol{X_0} (\boldsymbol{X_0}^t \boldsymbol{X_0})^{-1} \boldsymbol{X_0}^t \right) (\boldsymbol{X_1 b_1}) \right] \\ &= \frac{1}{\sigma^2} \| (\boldsymbol{I_n} - \boldsymbol{P}_{\boldsymbol{X}_0}) \boldsymbol{X}_1 \boldsymbol{b_1} \|^2 \end{aligned}$$

Where $\boldsymbol{P}_{\boldsymbol{X}_0} = \boldsymbol{X}_0 \left( \boldsymbol{X}_0^t \boldsymbol{X}_0 \right)^{-1} \boldsymbol{X}_0^t$ is the projection matrix.

#### 2 Proof of Lemma 1

*Proof.* Consider the linear model $\boldsymbol{y} = \boldsymbol{X_0 b_0} + \boldsymbol{X_1 b_1} + \varepsilon$ and assume the errors follow a symmetric distribution with mean $\mathbf{0}$ and known variance $\sigma^2 \boldsymbol{I_n}$. Let the data $\boldsymbol{y}$ be generated under the alternative hypothesis in

$$\begin{aligned} &H_0 : \text{Effect size of } \boldsymbol{X_1} = 0 \quad \text{vs} \\ &H_1 : \text{Effect size of } \boldsymbol{X_1} = \delta \end{aligned} \tag{3}$$

Denote, $\hat{\boldsymbol{b}}_{\boldsymbol{0},\boldsymbol{H_1}}$ and $\hat{\boldsymbol{b}}_{\boldsymbol{1},\boldsymbol{H_1}}$ as the MLEs for model under $H_1$ and $\hat{\boldsymbol{b}}_{\boldsymbol{0},\boldsymbol{H_0}}$ be the MLE for model under $H_0$. Let $\boldsymbol{P_{X_0}}$ be the projection matrix of $\boldsymbol{X_0}$ and tr(.) denotes the trace of a matrix.

$$\begin{aligned} E(\boldsymbol{y}|H_1) &= \boldsymbol{X_0 b_0} + \boldsymbol{X_1 b_1} \\ \hat{\boldsymbol{b}}_{\boldsymbol{0},\boldsymbol{H_0}} &= (\boldsymbol{X_0^t X_0})^{-1} \boldsymbol{X_0^t y} \\ \hat{\boldsymbol{b}}_{\boldsymbol{1},\boldsymbol{H_1}} &= \left\{ \boldsymbol{X_1^t} \left( \boldsymbol{I_n} - \boldsymbol{P_{X_0}} \right) \boldsymbol{X_1} \right\}^{-1} \boldsymbol{X_1^t} \left( \boldsymbol{I_n} - \boldsymbol{P_{X_0}} \right) \boldsymbol{y} \\ \hat{\boldsymbol{b}}_{\boldsymbol{0},\boldsymbol{H_1}} &= (\boldsymbol{X_0^t X_0})^{-1} \boldsymbol{X_0^t} \left( \boldsymbol{y} - \boldsymbol{X_1} \hat{\boldsymbol{b}}_{\boldsymbol{1},\boldsymbol{H_1}} \right) \\ \boldsymbol{P_{X_0}} &= \boldsymbol{X}_0 (\boldsymbol{X_0^t X_0})^{-1} \boldsymbol{X_0^t} \end{aligned}$$

$$f^2 = \left\{ \frac{R^2{}_{\boldsymbol{y},\boldsymbol{X_0},\boldsymbol{X_1}} - R^2{}_{\boldsymbol{y},\boldsymbol{X_0}}}{1 - R^2{}_{\boldsymbol{y},\boldsymbol{X}_0,\boldsymbol{X_1}}} \right\} = \frac{\left\{ \boldsymbol{X_0} \hat{\boldsymbol{b}}_{\boldsymbol{0},\boldsymbol{H_1}} + \boldsymbol{X_1} \hat{\boldsymbol{b}}_{\boldsymbol{1},\boldsymbol{H_1}} \right\}^t \boldsymbol{y} - \left( \boldsymbol{X_0} \hat{\boldsymbol{b}}_{\boldsymbol{0},\boldsymbol{H_0}} \right)^t \boldsymbol{y}}{\boldsymbol{y}^t \boldsymbol{y} - \left\{ \boldsymbol{X_0} \hat{\boldsymbol{b}}_{\boldsymbol{0},\boldsymbol{H_1}} + \boldsymbol{X_1} \hat{\boldsymbol{b}}_{\boldsymbol{1},\boldsymbol{H_1}} \right\}^t \boldsymbol{y}} = \frac{\boldsymbol{y}^t \boldsymbol{A} \boldsymbol{y}}{\boldsymbol{y}^t \boldsymbol{B} \boldsymbol{y}}, \tag{4}$$

where

$$\begin{aligned} \boldsymbol{A} &= \left( \boldsymbol{I_n} - \boldsymbol{P_{X_0}} \right) \boldsymbol{X_1} \left\{ \boldsymbol{X_1^t} \left( \boldsymbol{I_n} - \boldsymbol{P_{X_0}} \right) \boldsymbol{X_1} \right\}^{-1} \boldsymbol{X_1^t} \left( \boldsymbol{I_n} - \boldsymbol{P_{X_0}} \right) \\ \boldsymbol{B} &= \left\{ \left( \boldsymbol{I_n} - \boldsymbol{P_{X_0}} \right) - \boldsymbol{A} \right\}. \end{aligned}$$

Firstly note that, $\hat{f}^2$ in expression (4) can be expressed as a ratio of quadratic forms with $\boldsymbol{y}^t \boldsymbol{A} \boldsymbol{y} > 0$ and $\boldsymbol{y}^t \boldsymbol{B} \boldsymbol{y} > 0$. Secondly, both $\boldsymbol{A}$ and $\boldsymbol{B}$ are idempotent matrices with $\boldsymbol{A}^2 = \boldsymbol{A}$, $\boldsymbol{B}^2 = \boldsymbol{B}$ and $\boldsymbol{AB} = \mathbf{0}$.

Therefore we have the following:

$$\begin{aligned}
E\left(\boldsymbol{y}^t\boldsymbol{A}\boldsymbol{y}\right) &= \sigma^2\text{tr}(\boldsymbol{A}) + \boldsymbol{\mu}^t\boldsymbol{A}\boldsymbol{\mu} = \sigma^2 p_1 + \|\left(\boldsymbol{I_n} - \boldsymbol{P_{X_0}}\right)\boldsymbol{X_1}\boldsymbol{b_{1,H_1}}\|^2 \\
E\left(\boldsymbol{y}^t\boldsymbol{B}\boldsymbol{y}\right) &= \sigma^2\text{tr}(\boldsymbol{B}) + \boldsymbol{\mu}^t\boldsymbol{B}\boldsymbol{\mu} = \sigma^2\left(n - p_0 - p_1\right) \quad [\text{since}, \boldsymbol{\mu}^t\boldsymbol{B}\boldsymbol{\mu} = 0] \\
\text{var}\left(\boldsymbol{y}^t\boldsymbol{A}\boldsymbol{y}\right) &= 2\sigma^4\text{tr}(\boldsymbol{A}^2) + 4\sigma^2\boldsymbol{\mu}^t\boldsymbol{A}^2\boldsymbol{\mu} = 2\sigma^4 p_1 + 4\sigma^2\|\left(\boldsymbol{I_n} - \boldsymbol{P_{X_0}}\right)\boldsymbol{X_1}\boldsymbol{b_{1,H_1}}\|^2 \\
\text{var}\left(\boldsymbol{y}^t\boldsymbol{B}\boldsymbol{y}\right) &= 2\sigma^4\text{tr}(\boldsymbol{B}^2) + 4\sigma^2\boldsymbol{\mu}^t\boldsymbol{B}^2\boldsymbol{\mu} = 2\sigma^4(n - p_0 - p_1) \\
\text{cov}(\boldsymbol{y}^t\boldsymbol{A}\boldsymbol{y}, \boldsymbol{y}^t\boldsymbol{B}\boldsymbol{y}) &= 2\sigma^4\text{tr}(\boldsymbol{A}\boldsymbol{B}) + 4\sigma^2\boldsymbol{\mu}^t\boldsymbol{A}\boldsymbol{B}\boldsymbol{\mu} = 0
\end{aligned}$$

Using a second order Taylor series expansion on $\frac{\boldsymbol{y}^t\boldsymbol{A}\boldsymbol{y}}{\boldsymbol{y}^t\boldsymbol{B}\boldsymbol{y}}$ around its mean, we get:

$$\begin{aligned}
E\left(\frac{\boldsymbol{y}^t\boldsymbol{A}\boldsymbol{y}}{\boldsymbol{y}^t\boldsymbol{B}\boldsymbol{y}}\right) &\approx \frac{E\left(\boldsymbol{y}^t\boldsymbol{A}\boldsymbol{y}\right)}{E\left(\boldsymbol{y}^t\boldsymbol{B}\boldsymbol{y}\right)} - \frac{\text{cov}(\boldsymbol{y}^t\boldsymbol{A}\boldsymbol{y}, \boldsymbol{y}^t\boldsymbol{B}\boldsymbol{y})}{\{E\left(\boldsymbol{y}^t\boldsymbol{B}\boldsymbol{y}\right)\}^2} + \frac{\text{var}\left(\boldsymbol{y}^t\boldsymbol{B}\boldsymbol{y}\right)E\left(\boldsymbol{y}^t\boldsymbol{A}\boldsymbol{y}\right)}{\{E\left(\boldsymbol{y}^t\boldsymbol{B}\boldsymbol{y}\right)\}^3} \\
&= \frac{\|\left(\boldsymbol{I_n} - \boldsymbol{P_{X_0}}\right)\boldsymbol{X_1}\boldsymbol{b_1}\|^2}{n\sigma^2} + o(1), \quad \text{as } n \to \infty
\end{aligned} \tag{5}$$

To obtain an expression for $\text{var}\left(\frac{\boldsymbol{y}^t\boldsymbol{A}\boldsymbol{y}}{\boldsymbol{y}^t\boldsymbol{B}\boldsymbol{y}}\right)$, we have used a first order Taylor series expansion.

$$\begin{aligned}
\text{var}\left(\frac{\boldsymbol{y}^t\boldsymbol{A}\boldsymbol{y}}{\boldsymbol{y}^t\boldsymbol{B}\boldsymbol{y}}\right) &\approx \frac{\{E(\boldsymbol{y}^t\boldsymbol{A}\boldsymbol{y})\}^2}{\{E(\boldsymbol{y}^t\boldsymbol{B}\boldsymbol{y})\}^2}\left[\frac{\text{var}(\boldsymbol{y}^t\boldsymbol{A}\boldsymbol{y})}{\{E(\boldsymbol{y}^t\boldsymbol{A}\boldsymbol{y})\}^2} - \frac{2\text{cov}(\boldsymbol{y}^t\boldsymbol{A}\boldsymbol{y}, \boldsymbol{y}^t\boldsymbol{B}\boldsymbol{y})}{E(\boldsymbol{y}^t\boldsymbol{A}\boldsymbol{y})E(\boldsymbol{y}^t\boldsymbol{B}\boldsymbol{y})} + \frac{\text{var}(\boldsymbol{y}^t\boldsymbol{B}\boldsymbol{y})}{\{E(\boldsymbol{y}^t\boldsymbol{B}\boldsymbol{y})\}^2}\right] \\
&\to 0, \quad \text{as } n \to \infty
\end{aligned} \tag{6}$$

Using Chebyshev's inequality and combining expressions (5) and (6), we arrive at the main result. □

#### 3 Cohen's $f^2$ in Generalized Linear Models

Now consider a generalized linear model (McCullagh and Nelder (1989)). Let, the outcome variable $\boldsymbol{y}$ follows the exponential family, $\exp[\{\boldsymbol{y}\theta - b(\theta)\}/a(\phi) + c(\boldsymbol{y}, \phi)]$. Let $\mu = E(\boldsymbol{y})$ and $g(.)$ be the link function. $\mu$ is related to the canonical parameter $\theta$ through the function $\mu = b'(\theta)$, where $b'$ denotes the first derivative of $b$. Then the model is completed by $\eta = g(\mu) = \boldsymbol{X_0}\boldsymbol{b_0} + \boldsymbol{X_1}\boldsymbol{b_1}$. Here the standard test is likelihood ratio test for testing of hypothesis, $\Lambda(\boldsymbol{y}) = 2\left\{\ell(\hat{\boldsymbol{b}}_{\boldsymbol{1},\boldsymbol{H_1}}, \hat{\boldsymbol{b}}_{\boldsymbol{0},\boldsymbol{H_1}}|\boldsymbol{X_0}, \boldsymbol{X_1}) - \ell(\hat{\boldsymbol{b}}_{\boldsymbol{0},\boldsymbol{H_0}}|\boldsymbol{X_0})\right\}$. As the sample size $n \to \infty$, the likelihood ratio statistic $\Lambda(\boldsymbol{y})$ follows a central chi-squared distribution $\chi^2_{p_1}$ with $p_1$ degrees of freedom, when $\boldsymbol{y}$ is generated under the model in $H_0$. $\Lambda(\boldsymbol{y})$ follows a non-central chi-squared distribution $\chi^2_{p_1}(\gamma_n)$ with degrees of freedom $p_1$ and non-centrality parameter $\gamma_n$, when $\boldsymbol{y}$ is generated under $H_1$. However, there is no simple and explicit form for $\gamma_n$. Self et al. (1992) and Shieh (2000) defined $\gamma_n$ in likelihood ratio test for generalized linear models as $\gamma_n := E_{\boldsymbol{y}\sim H_1}\{\Lambda(\boldsymbol{y})\}$. Similar to the non-centrality parameter in linear regression, $\gamma_n$ grows linearly with $n$ since $E_{\boldsymbol{y}\sim H_1}\{\Lambda(\boldsymbol{y})\} = n\Big\{2a^{-1}(\phi)\,\{b'(\theta)[\theta-\theta^*] - [b(\theta) - b(\theta^*)]\}\Big\} + o(1)$ (where $\theta$ and $\theta^*$ denote the canonical parameter values evaluated at $(\boldsymbol{b_0}, \boldsymbol{b_1})$ and $(\boldsymbol{b_0^*}, \boldsymbol{b_1} = \boldsymbol{0})$ and $\boldsymbol{b_0^*}$ is the limiting value of $\hat{\boldsymbol{b}}_{\boldsymbol{0},\boldsymbol{H_0}}$ as described in equation (2.2) of Self and Mauritsen (1988) (see Cordeiro (1983) and section 2 of Shieh (2000) for a detailed derivation)). Consider the adjusted coefficient of determination for generalized linear models as shown below (note that the definition of squared multiple correlation is generally accepted for linear regression but it is not directly applied to a generalized linear models (for example a logistic regression) and there remains a lack of general consensus discussed in literature (see Liao and McGee (2003) for more details)), $R^2_l = 1 - \frac{\ell(\boldsymbol{y}|\text{ Any predictor }\boldsymbol{X})}{\ell(\boldsymbol{y}|\text{ Intercept Only Model})}$. Then Cohen's $f^2$ in generalized linear models can be written as,

$$f^2 = 2\left\{\frac{R^2_{l,H_1} - R^2_{l,H_0}}{1 - R^2_{l,H_1}}\right\} := 2\left\{\frac{\ell(\boldsymbol{b_{1,H_1}}, \boldsymbol{b_{0,H_1}}|\boldsymbol{X_0}, \boldsymbol{X_1}) - \ell(\boldsymbol{b_{0,H_0}}|\boldsymbol{X_0})}{\ell(\boldsymbol{b_{1,H_1}}, \boldsymbol{b_{0,H_1}}|\boldsymbol{X_0}, \boldsymbol{X_1})}\right\}, \tag{7}$$

where, $R^2_{l,H_0}$ and $R^2_{l,H_1}$ are the adjusted coefficient of determinations under the null and the alternative respectively.

#### 3.1 Connection between Cohen's $f^2$ and non-centrality parameter in Generalized Linear Models

First denote, $\hat{\ell}_{H_1} := \ell(\hat{\boldsymbol{b}}_{\boldsymbol{1},\boldsymbol{H_1}}, \hat{\boldsymbol{b}}_{\boldsymbol{0},\boldsymbol{H_1}}|\boldsymbol{X_0}, \boldsymbol{X_1})$ and $\hat{\ell}_{H_0} := \ell(\hat{\boldsymbol{b}}_{\boldsymbol{0},\boldsymbol{H_0}}|\boldsymbol{X_0})$. Then,

$$f^2 = 2\left\{\frac{\hat{\ell}_{H_1} - \hat{\ell}_{H_0}}{\hat{\ell}_{H_1}}\right\}$$

Similar to the proof of Lemma 1, we will use Taylor series expansion around the mean and variance. To derive the $E_{\boldsymbol{y}\sim H_1}\{\hat{\ell}_{H_1}\}$, note that in the generalized linear model,

$$\ell_{H_1} = \sum_{i=1}^{n} \{(\theta y_i - b(\theta))/a(\phi) + c(y_i, \phi)\}.$$

Hence, $E_{\boldsymbol{y}\sim H_1}\{\ell_{H_1}\} = O(n)$. Moreover following Lawley (1956) we have,

$$\begin{aligned} E_{\boldsymbol{y}\sim H_1}\{\hat{\ell}_{H_1}\} &= E_{\boldsymbol{y}\sim H_1}\left\{\ell_{H_1} + \hat{\ell}_{H_1} - \ell_{H_1}\right\} \\ &= O(n) + \frac{1}{2}\{p_0 + p_1 + O(1/n) + O(1/n^2)\} \end{aligned}$$

Similarly, $E_{\boldsymbol{y}\sim H_1}\{2(\hat{\ell}_{H_1} - \hat{\ell}_{H_0})\} = \gamma_n$, $\text{Var}_{\boldsymbol{y}\sim H_1}\{2(\hat{\ell}_{H_1} - \hat{\ell}_{H_0})\} = O(n)$ and $\text{Var}_{\boldsymbol{y}\sim H_1}\{\hat{\ell}_{H_1}\} = O(n)$ (see Lawley (1956) and Cordeiro (1983) for exact expressions). Next, using the Taylor series expansions specified in (5) and (6), we have

$$\begin{aligned} E_{\boldsymbol{y}\sim H_1}\left\{f^2\right\} &= \frac{\gamma_n}{O(n)} + o(1), \quad \text{as} \quad n \to \infty \\ \text{Var}_{\boldsymbol{y}\sim H_1}\left\{f^2\right\} &\to 0, \quad \text{as} \quad n \to \infty \end{aligned}$$

### 4 Proof of Theorem 1

#### 4.1 Expression of T

*Proof.* Define, $z(n) = \frac{T - p_1 - n\delta}{\sqrt{2(p_1 + 2n\delta)}}$. Consider the following equation for type 1 and type 2 errors,

$$P\left(\chi^2_{p_1} > \text{T}|\gamma_n = 0\right) = P\left(\chi^2_{p_1}(\gamma_n) < \text{T}|\gamma_n = n\delta\right). \tag{8}$$

Further, in case of the non-central chi-square distribution, we assume that the normality approximation holds true.

$$P\left(\chi^2_{p_1} > \text{T}|\gamma_n = 0\right) = \Phi\left(\frac{\text{T} - p_1 - n\delta}{\sqrt{2(p_1 + 2n\delta)}}\right) + o(1). \tag{9}$$

Since (9) is true for every $n$, taking a derivative of (9) with respect to $n$ we get,

$$\frac{d}{dn}\mathrm{P}(\chi^2_{p_1} > T) = -\frac{T^{\frac{p_1}{2}-1}\exp(-T/2)}{2^{\frac{p_1}{2}}\Gamma(\frac{p_1}{2})}\frac{dT}{dn}$$
$$\frac{d}{dn}\Phi\left(\frac{T-p_1-n\delta}{\sqrt{2(p_1+2n\delta)}}\right) = \frac{1}{\sqrt{2\pi}}\exp\left(-\frac{z^2(n)}{2}\right)\frac{dz(n)}{dn}$$
$$\frac{dz(n)}{dn} = \frac{\left(\frac{dT}{dn}-\delta\right)}{\sqrt{2(p_1+2n\delta)}} - \frac{2\delta\,(T-p_1-n\delta)}{\{2(p_1+2n\delta)\}^{3/2}}$$

After equating the derivatives and some readjustment of terms, we get

$$\frac{dT}{dn} = \frac{\delta\left\{1+\sqrt{2(p_1+2n\delta)}b\right\}}{1+\sqrt{2(p_1+2n\delta)}a} \tag{10}$$

with $a = \left\{\frac{1}{\sqrt{2\pi}}\exp\left(-\frac{z^2(n)}{2}\right)\right\}^{-1}\frac{T^{\frac{p_1}{2}-1}\exp(-T/2)}{2^{\frac{p_1}{2}}\Gamma(\frac{p_1}{2})}$ and $b = \frac{2z(n)}{2(p_1+2n\delta)}$.

*Assumption C.1* In order to derive an expression for $T$, we will first assume that $1+\sqrt{2(p_1+2n\delta)}a = K(\text{say})$ is asymptotically constant with $K > 1$.

Later we will show indeed $K \to 2+\sqrt{2}$ as $n \to \infty$. From (10), we further obtain,

$$\frac{K}{\delta}\frac{dT}{dn} = \frac{T+n\delta}{2n\delta}\left(1+o(1)\right)$$

Solving the first order differential equation while ignoring the $o(1)$ term, we derive the expression for $T$,

$$T = \frac{\delta n}{2K-1} + cn^{\frac{1}{2K}}, \quad \text{where, } c \text{ is a constant of integration} \tag{11}$$

Now, we will focus on the asymptotics of $K$.

$$\begin{aligned} K &= 1+\sqrt{2(p_1+2n\delta)}a \\ &= 1+\exp\left\{\log\sqrt{2(p_1+2n\delta)}+\log(a)\right\} \\ &= 1+\exp\left\{\frac{1}{2}\log\{2p_1+4n\delta\}+\left(\frac{p_1}{2}-1\right)\log(T)-\frac{T}{2}+\frac{(T-p_1-n\delta)^2}{4(p_1+2n\delta)}+k_1\right\} \end{aligned} \tag{12}$$

where, $k_1 = -\log\{2^{\frac{p_1}{2}}\Gamma(p_1/2)\} + \log\sqrt{2\pi}$. Now we will expand each term in (12) to obtain an expression for $K$. We will assume, $\| \frac{p_1}{2n\delta} \| < 1$ and $\| \frac{c(2K-1)}{\delta n^{1-\frac{1}{2K}}} \| < 1$ which is true for $p_1 << n$ and particularly when $p_1 = 1$ in exposure-mixture models. Separately expanding the terms in (12), we get:

$$\begin{aligned} \frac{1}{2}\log\{2p_1 + 4n\delta\} &= \frac{\log(n)}{2} + \frac{\log(4\delta)}{2} + o(1) \\ \left(\frac{p_1}{2} - 1\right)\log(T) &= \left(\frac{p_1}{2} - 1\right)\log(n) + \left(\frac{p_1}{2} - 1\right)\log\left(\frac{\delta}{2K-1}\right) + o(1) \\ \frac{(T - p_1 - n\delta)^2}{4(p_1 + 2n\delta)} &= \frac{n\delta}{8(2K-1)^2} + \frac{cn^{\frac{1}{2K}}}{4(2K-1)} - \frac{T}{4} + \frac{n\delta}{8} + o(1) \end{aligned} \tag{13}$$

Now putting the expressions in (13) in the final expression of (12), we get

$$\begin{aligned} K &= 1 + \exp\left\{N - \frac{3T}{4} + k_1 + o(1)\right\} \\ \text{where, } N &= \frac{n\delta}{8(2K-1)^2} + \frac{cn^{\frac{1}{2K}}}{4(2K-1)} + \frac{n\delta}{8} + \left(\frac{p_1 - 1}{2}\right)\log(n) \end{aligned} \tag{14}$$

Note that $N \to \infty$ as $n \to \infty$. Separately using the expressions of $T$ in (11) and $N$ in (14), asymptotically we obtain a expression for $\frac{T}{N}$,

$$\frac{3T/4}{N} = \frac{3(2K-1)}{2K^2 - 2K + 1} + o(1) \tag{15}$$

Then putting the expression in (15) in (14), $K$ can be rewritten as,

$$K = 1 + \exp\left\{N\left(1 - \frac{3(2K-1)}{2K^2 - 2K + 1} + o(1)\right) + k_1 + o(1)\right\} \tag{16}$$

which after some rearrangement of terms can be rewritten as,

$$\frac{\log(K-1)}{N} = \left\{1 - \frac{3(2K-1)}{2K^2 - 2K + 1} + o(1)\right\} + \frac{k_1 + o(1)}{N}$$

Under the assumption that $K$ is asymptotically a constant (Assumption C.1), we note that both the terms $\frac{\log(K-1)}{N}$ and $\frac{k_1 + o(1)}{N}$ tend to 0, as $n \to \infty$. Therefore as $n \to \infty$, ignoring the $o(1)$ term, the remaining $\left\{1 - \frac{3(2K-1)}{2K^2-2K+1}\right\}$ must also tend to 0. Solving the quadratic equation, we obtain, $\big(K \to 2 + \sqrt{2}\big)(1 + o(1)\big)$.

Therefore, the expression of $T$ can be rewritten as,

$$T = \left( \frac{\delta n}{2K - 1} + cn^{\frac{1}{2K}} \right) (1 + o(1)),$$

with $\big(K \to 2 + \sqrt{2})(1 + o(1)\big)$ and $c$ being the constant of integration. □

#### 4.2 Expression of $\log \alpha$ and $\log \beta$

*Proof.* Consider the left hand side of equation (8) (which is the expression of type 1 error $\alpha(\mathrm{T})$),

$$P\left(\chi^2_{p_1} > \mathrm{T} | \gamma_n = 0\right) = \Phi\left(\frac{p_1 - T}{\sqrt{2p_1}}\right)$$

$T \to \infty$ as $n \to \infty$. Therefore, the type 1 error goes to zero for large $n$. Since we have constrained $\alpha(T) = \beta(T)$, the type 2 error also goes to zero. Now, $\forall x \in R$, define the erf as,

$$\Phi(x) = \frac{1}{\sqrt{2\pi}} \int_x^{-\infty} e^{\frac{-t^2}{2}} dt = \frac{1}{2}\left[1 + \mathrm{erf}\left(\frac{x}{\sqrt{2}}\right)\right]$$

We will particularly use an approximation obtained by Hans Heinrich Bürmann's theorem (Schöpf and Supancic (2014)), which converges more rapidly for all real valued $x$ than a Taylor series expansion. For all real $x$, the Bürmann series is given below:

$$\mathrm{erf}(x) \approx \frac{2}{\sqrt{\pi}} \mathrm{sgn}(x) \sqrt{1 - e^{-x^2}} \left( \frac{\sqrt{\pi}}{2} + \frac{31}{200} e^{-x^2} - \frac{341}{8000} e^{-2x^2} \right)$$

When $x$ is large, we could ignore the terms containing $e^{-x^2}$ and $e^{-2x^2}$. Therefore, when $x \to \infty$, we have,

$$\mathrm{erf}(x) = \mathrm{sgn}(x) \sqrt{1 - e^{-x^2}}\,(1 + o(1))$$

When the type 1 and type 2 errors for Calibrated Cutoff goes to zero, $x$ in the argument of $\Phi(x)$, goes to $-\infty$. Therefore,$\Phi(x)$ can be approximated as,

$$\Phi(x) = \frac{1}{2}\left[1 - \sqrt{1 - \exp\left\{-\frac{x^2}{2}\right\}}\right](1 + o(1))$$

which after some algebra can be expressed as (ignoring the $\Phi^2(x)$ term),

$$\log\{\Phi(x)\} = \left\{-\frac{x^2}{2} - \log 4\right\}(1 + o(1)) \tag{17}$$

Now let's denote $x = z(n)$, which is defined as $z(n) = \frac{T - p_1 - n\delta}{\sqrt{2(p_1 + 2n\delta)}}$. Note that, $\Phi(z(n))$ is the type 2 error from (8). Using (17), we get

$$\log\{\Phi(z(n))\} = -\frac{1}{2}\left\{\frac{T - p_1 - n\delta}{\sqrt{2(p_1 + 2n\delta)}}\right\}^2 - \log 4 + o(1)$$

Taking derivative with respect to $n$ on both the sides and using the expressions in (13),

$$\frac{d\log\{\Phi(z(n))\}}{dn} = -\frac{d}{dn}\left\{\frac{n\delta}{8(2K-1)^2} + \frac{cn^{\frac{1}{2K}}}{4(2K-1)} - \frac{T}{4} + \frac{n\delta}{8} - \log 4 + o(1)\right\} + o(1)$$

Finally we arrive at

$$\frac{d}{dn}\log\{\alpha(T)\} = \frac{d}{dn}\log\{\beta(T)\} = -\frac{\delta(K-1)^2}{2(2K-1)^2} + o(1)$$

The expressions of $\log\{\alpha(T)\}$ and $\log\{\beta(T)\}$ can be obtained by solving the first order differential equation and rearranging the constant terms. □

### 5 Calibrated type 1 and type 2 errors remain approximately same under T

We conducted Monte Carlo simulation using linear regression to estimate the calibrated type 1 and type 2 errors under the framework of T and hypothesis in $H_0 : f^2 = 0 \quad \text{vs} \quad H_1 : f^2 = \delta$ to substantiate results of Theorem 1. Five covariates were generated for the smaller baseline model with $R^2$ set at $20\%$. Two separate larger models were used - one with a single exposure and five baseline covariates and another with three exposures and five baseline covariates. The true Cohen's $f^2$ for the larger models were set at $2\%, 4\%$ and $6\%$ respectively. Each simulation was iterated at least $10^5$ times. The data generating procedure is elaborated in supplementary materials. For a fixed $n$, as the value of $f^2$ increases, the error calibrated cutoff T also increases and the corresponding the type 1 and type 2 errors decrease. The results presented in Table 1 confirm that the estimated type 1 and type 2 errors are approximately equal and the results get better as sample sizes increase.

### 6 Type 2 error function

Theorem 1 suffices when the data is generated under the Cohen's $f^2 = \delta$ and the alternative is set at $H_1 : f^2 = \delta$. But what happens when the true $f^2$, under which the data is generated, is not $\delta$ ? To highlight and investigate these subtleties, let us assume that the true Cohen's $f^2 = \epsilon$, under which the data is generated. Then given the hypothesis in (3), the type 2 error function is, $\beta(\epsilon) = P\left(\chi^2_{p_1}(\gamma_n) < \mathrm{T}(\delta) | \gamma_n = n\epsilon\right)$.

#### 6.1 Case 1: when $0 < \epsilon < \delta$.

Note that, $\beta(0) \to 1$ as $n \to \infty$. Since $1 - \beta(0) = \beta(\delta) \to 0$ when $n \to \infty$, there exists a point $\epsilon^* \in (0, \delta)$ such that $\beta(\epsilon) > 0.5$ for $\epsilon \in (0, \epsilon^*)$ and $\beta(\epsilon) \leq 0.5$ for $\epsilon \in [\epsilon^*, \delta)$. The $\epsilon^*$ is the root to $P\left(\chi^2_{p_1}(\gamma_n) < \mathrm{T}(\delta) | \gamma_n = n\epsilon^*\right) = \frac{1}{2}$. The following corollary introduces an asymptotic expression for $\epsilon^*$ under the hypothesis (3).

**Corollary 1.** *Under the hypothesis (3) and as* $n \to \infty$, $\epsilon^* = \frac{\delta}{2K-1} + o(1)$ *with* $K = (2 + \sqrt{2})(1 + o(1))$

#### 6.2 Case 2: when $\epsilon \geq \delta$.

First note that $\beta(\epsilon)$ is monotonically decreasing in $\epsilon$ (see sub-section S.6.4). Therefore the type 2 error is even smaller when the true Cohen's $f^2$ is larger than $\delta$. Hence asymptotically the $\beta(\epsilon) \to 0$ (see supplementary material). Now we investigate what happens to the type 2 error when $\boldsymbol{y}$ is generated while $\epsilon$ lies between $(0, \delta)$.

#### 6.3 Neutral effect size, null and alternative neighborhood.

When true Cohen's $f^2$, $\epsilon \in [0, \epsilon^*)$, T prefers the $H_0$ with probability greater than $\frac{1}{2}$. Therefore, we can think of the interval $[0, \epsilon^*)$ as an expanded null hypothesis which nicely connects to the interval null hypothesis discussed in literature (Morey and Rouder (2011), Kruschke (2013) and Liao et al. (2020), Midya and Liao (2021)). We name $\epsilon^*$ as the "neutral effect size". Note that, $\epsilon^*$ decreases as $n$ increases although very slowly due to a term $O\left(\frac{1}{n^{1-\frac{1}{2K}}}\right)$ and eventually converges to $\frac{\delta}{2K-1}$. The interpretation of "neutral effect size" is that, as long as the true Cohen's $f^2$, denoted by $\epsilon$, originates below $\epsilon^*$, the probability of rejecting the $H_0$ gets smaller than $\frac{1}{2}$ even when the true effect size $\epsilon$ is far from zero. Whereas, if true effect size $\epsilon$ originates above $\epsilon^*$, the probability of rejecting the null becomes greater than $\frac{1}{2}$. Similarly, an interval of the form $\{x|x > \epsilon^*\}$ can be conceived such that, for any $\epsilon > \epsilon^*$, the probability of rejecting the null remains greater than $\frac{1}{2}$. The interval denoted is named as the "alternative neighborhood".

To demonstrate the concepts discussed above, $\beta(\epsilon)$, i.e. the type 2 error function is plotted for $n = 250$ and $n = 1000$ at $p_1 = 5$ and $\delta = 10\%$ in Figure 1 with $\epsilon$ from 0 to $10\%$ on x-axis. $\beta(\epsilon)$ decreases smoothly starting from 0 as $\epsilon$ increases. The neutral effect sizes, $\epsilon^*$ are $2.6\%$ and $2.2\%$ for $n = 250$ and $n = 1000$ respectively. The shaded red region denotes the "alternative neighborhood" with type 2 error below $\frac{1}{2}$, whereas the shaded blue region denotes the "null neighborhood" with type 2 error above $\frac{1}{2}$. As long as the true effect size ($\epsilon$) of the underlying data, is greater than the neutral effect size, the error calibrated cutoff T will favour the alternative. Whereas the null will be favoured only when the true effect sizes are less than the neutral effect sizes. Note that, neutral effect size, $\epsilon^*$ decreases from $2.6\%$ to $2.2\%$ as the sample size $n$ increases but the rate of decrease is very slow. This plot therefore reinforces the idea that no matter how large the sample size is, this error calibrated cutoff will only reject null in the favour of the alternative if and only if the true effect size originates from a neighbourhood $\{f^2|f^2 > \epsilon^*\}$.

#### 6.4 $\beta(\epsilon)$ is monotonically decreasing in $\epsilon$

*Proof.* Similar to (9), we use the normality approximation of the non-central chi-square distribution for large $n$ as below,

$$\beta(\epsilon) = P\left(\chi^2_{p_1}(\gamma_n) < \mathrm{T}(\delta)|\gamma_n = n\epsilon\right) = \Phi\left(\frac{\mathrm{T}(\delta) - p_1 - n\epsilon}{\sqrt{2(p_1 + 2n\epsilon)}}\right)(1 + o(1))$$

Since $\Phi(.)$ is a non-decreasing function, it suffices to show that the argument $\frac{\mathrm{T}(\delta)-p_1-n\epsilon}{\sqrt{2(p_1+2n\epsilon)}}$ is monotonically decreasing in $\epsilon$. Now note that,

$$\frac{d}{d\epsilon}\left(\frac{\mathrm{T}(\delta)-p_1-n\epsilon}{\sqrt{2(p_1+2n\epsilon)}}\right)=\frac{-2n\mathrm{T}(\delta)-2n^2\epsilon}{\{2(p_1+2n\epsilon)\}^{3/2}}<0$$

□

### 7 Proof of Corollary 1

*Proof.* When $\mathrm{T}(\delta^*)=\Lambda(\boldsymbol{y})$, $\delta$ also attains its maximum since $\mathrm{T}(\delta)$ is an monotonically increasing function of $\delta$. Using the expression of $\mathrm{T}(\delta)$ from Theorem 1 and solving the equation, we get a closed form expression for $\delta^*$,

$$\delta^*=\frac{\Lambda(\boldsymbol{y})-c_1 n^{\frac{1}{2K}}}{n}(2K-1)+o(1)$$

As $n\to\infty$, both the type 1 and type 2 errors can be approximated by standard normal cdfs, $\Phi\left(\frac{p_1-\mathrm{T}(\delta^*+h)}{\sqrt{2p_1}}\right)$ and $\Phi\left(\frac{\mathrm{T}(\delta^*+h)-p_1-E_{\boldsymbol{y}}\{\Lambda(\boldsymbol{y})\}}{\sqrt{2(p_1+2E_{\boldsymbol{y}}\{\Lambda(\boldsymbol{y})\})}}\right)$ respectively for $h\geq-\delta^*$. The proof is completed by noting that $\mathrm{T}(\delta^*+h)$ is a monotonically increasing function of $h$. □

### 8 Data generating process for simulations

We followed the setup elaborated in Shen and Ye (2002) and George and Foster (2000) but modified some key steps as noted below.

1. First, we generated design matrices, $\boldsymbol{X_0}$ and $\boldsymbol{X_1}$ with dimensions $n\times p_0$ and $n\times(p_0+p_1)$ respectively. The covariates were sampled from $\mathcal{N}(\mathbf{0},\Sigma_k^{(p_k)\times(p_k)}), k=0,1$ where the $(i,j)^{th}$ entry of $\Sigma_k$ had the form $\rho+\psi$ with $\psi\sim\mathcal{N}(0.1,0.01)$. We assumed the error follows a normal distribution with mean zero and known variance 1. This was intentionally done to keep the task of estimating unknown variance and hypothesis testing separate. We choose $\rho=0.6$ to reflect high correlation within a group of exposures, which is usually observed in chemical exposure analyses. To quantify the contribution/impact of the two-sets of predictors, we assumed that $\boldsymbol{X_0}$ and $\boldsymbol{X_1}$ are orthogonal, or $\boldsymbol{X_1}$ can be

orthogonalized by the transformation $(\boldsymbol{I_n} - \boldsymbol{X_0}(\boldsymbol{X_0^t X_0})^{-1}\boldsymbol{X_0^t})\boldsymbol{X_1}$.

2. Under $H_0$, the population level coefficient of variation is denoted by $\boldsymbol{R^2_{y,X_0}}$ and the corresponding estimate is $E_{\boldsymbol{y}\sim\boldsymbol{H}_0}\{\boldsymbol{R^2_{y,X_0}}\}$. A first order Taylor series expansion was used to obtain an expression of $E_{\boldsymbol{y}\sim\boldsymbol{H}_0}\{\boldsymbol{R^2_{y,X_0}}\}$ in terms of the beta coefficient $\boldsymbol{b_0}$ (18). Given the values of $E_{\boldsymbol{y}\sim\boldsymbol{H}_0}\{\boldsymbol{R^2_{y,X_0}}\}$, $n, p_0, \boldsymbol{X_0}, \boldsymbol{X_1}$ and known $\sigma$, the aim was to find the corresponding values of $\boldsymbol{b_0}$. For simplicity, we assumed that all the regression coefficients in $\boldsymbol{b_0}$ were equal.

$$E_{\boldsymbol{y}\sim\boldsymbol{H}_0}\{\boldsymbol{R^2_{y,X_0}}\} = E_{\boldsymbol{y}\sim\boldsymbol{H}_0}\left\{\frac{(\boldsymbol{X_0}\hat{\boldsymbol{b}}_0)^t\boldsymbol{y} - \frac{1}{n}\boldsymbol{y}^t\boldsymbol{1}\boldsymbol{1}^t\boldsymbol{y}}{\boldsymbol{y}^t\boldsymbol{y} - \frac{1}{n}\boldsymbol{y}^t\boldsymbol{1}\boldsymbol{1}^t\boldsymbol{y}}\right\} = \frac{\sigma^2(p_0-1) + \boldsymbol{b_0^t X_0^t}(\boldsymbol{I_n} - \frac{\boldsymbol{11}^t}{n})\boldsymbol{X_0 b_0}}{(n-1)\sigma^2 + \boldsymbol{b_0^t X_0^t}(\boldsymbol{I_n} - \frac{\boldsymbol{11}^t}{n})\boldsymbol{X_0 b_0}} + o(1), \tag{18}$$

Then $\boldsymbol{y}$ under $H_0$ was generated using a sum of $\boldsymbol{X_0 b_0}$ and normally distributed errors (with mean at zero and variance at one) with prespecified that $E_{\boldsymbol{y}\sim\boldsymbol{H}_0}\{\boldsymbol{R^2_{y,X_0}}\}$.

$$\boldsymbol{y}|\boldsymbol{H_0} = \boldsymbol{X_0 b_0} + \epsilon, \quad \epsilon \sim \mathcal{N}(0,1).$$

3. Under $H_1$, a population level $\boldsymbol{R^2_{y,X_0,X_1}}$ show the variability explained by both $\boldsymbol{X_0}$ and $\boldsymbol{X_1}$. Therefore, to obtain the values of $\boldsymbol{b_1}$ for generating $\boldsymbol{y}$ under the alternative, we first approximate an expression of $E_{\boldsymbol{y}\sim\boldsymbol{H}_1}\{\boldsymbol{R^2_{y,X_0,X_1}}\}$ (19). Once $E_{\boldsymbol{y}\sim\boldsymbol{H}_1}\{\boldsymbol{R^2_{y,X_0,X_1}}\}$ was fixed, the values of $\boldsymbol{b_1}$ were calculated using known $\sigma$, fixed $n, p_0, p_1, \boldsymbol{X_0}, \boldsymbol{X_1}$ and $\boldsymbol{b_0}$ obtained from step 2. For simplicity, we assumed all the regression coefficients in $\boldsymbol{b_1}$ are equal.

$$E_{\boldsymbol{y}\sim\boldsymbol{H}_1}\{\boldsymbol{R^2_{y,X_0,X_1}}\} = E_{\boldsymbol{y}\sim\boldsymbol{H}_1}\left\{\frac{(\boldsymbol{X_0}\hat{\boldsymbol{b}}_0 + \boldsymbol{X_1}\hat{\boldsymbol{b}}_1)^t\boldsymbol{y} - \frac{1}{n}\boldsymbol{y}^t\boldsymbol{1}\boldsymbol{1}^t\boldsymbol{y}}{\boldsymbol{y}^t\boldsymbol{y} - \frac{1}{n}\boldsymbol{y}^t\boldsymbol{1}\boldsymbol{1}^t\boldsymbol{y}}\right\} = \frac{\sigma^2(p_0+p_1-1)+(\boldsymbol{X_0 b_0}+\boldsymbol{X_1 b_1})^t(\boldsymbol{I_n}-\frac{\boldsymbol{11}^t}{n})(\boldsymbol{X_0 b_0}+\boldsymbol{X_1 b_1})}{(n-1)\sigma^2+(\boldsymbol{X_0 b_0}+\boldsymbol{X_1 b_1})^t(\boldsymbol{I_n}-\frac{\boldsymbol{11}^t}{n})(\boldsymbol{X_0 b_0}+\boldsymbol{X_1 b_1})} + o(1). \tag{19}$$

Then $\boldsymbol{y}$ under $H_1$ is generated using design matrices $\boldsymbol{X_0}$ and $\boldsymbol{X_1}$ with corresponding coefficients $\boldsymbol{b_0}$

and $\boldsymbol{b_1}$ obtained from step 2 and 3 such that $E_{\boldsymbol{y}\sim\boldsymbol{H_0}}\{\boldsymbol{R^2_{y,X_0}}\}$ and $E_{\boldsymbol{y}\sim\boldsymbol{H_1}}\{\boldsymbol{R^2_{y,X_0,X_1}}\}$ are prespecified. We assumed a normally distributed error with mean zero and variance one.

$$\boldsymbol{y}|\boldsymbol{H_1} = \boldsymbol{X_0}\boldsymbol{b_0} + \boldsymbol{X_1}\boldsymbol{b_1} + \epsilon, \quad \epsilon \sim \mathcal{N}(0,1).$$

### 9 PFAS and metal exposures and Serum lipoprotein-cholesterols outcomes

Serum PFAS, included in the study were Perfluorodecanoic acid (PFDeA), Perfluorohexane sulfonic acid (PFHxS), 2-(N-methylperfluoroctanesulfonamido)acetic acid (Me-PFOSA-AcOH), Perfluorononanoic acid (PFNA), Perfluoroundecanoic acid (PFUA); Perfluorododecanoic acid (PFDoA), n-perfluorooctanoic acid (n-PFOA), Branch perfluorooctanoic acid isomers (Sb-PFOA), n-perfluorooctane sulfonic acid (n-PFOS), Perfluoromethylheptane sulfonic acid isomers (Sm-PFOS). All PFAS had at least $38\%$ observations above the lower limit of detection (LLOD).

Metals included were Lead (Pb), Cadmium (Cd), Total Mercury (THg), Selenium (Se), Manganese (Mn) and were measured in who;e blood. All metals had at least $85\%$ observations above LLOD. See laboratory procedural manuals for further details. As the outcomes, we have chosen only two lipoprotein-cholesterols, serum low-density lipoprotein-cholesterols (LDL-C) and high-density lipoprotein-cholesterols (HDL-C) because of their differing roles in atherogenic dyslipidemia (Rohatgi et al. (2021)).

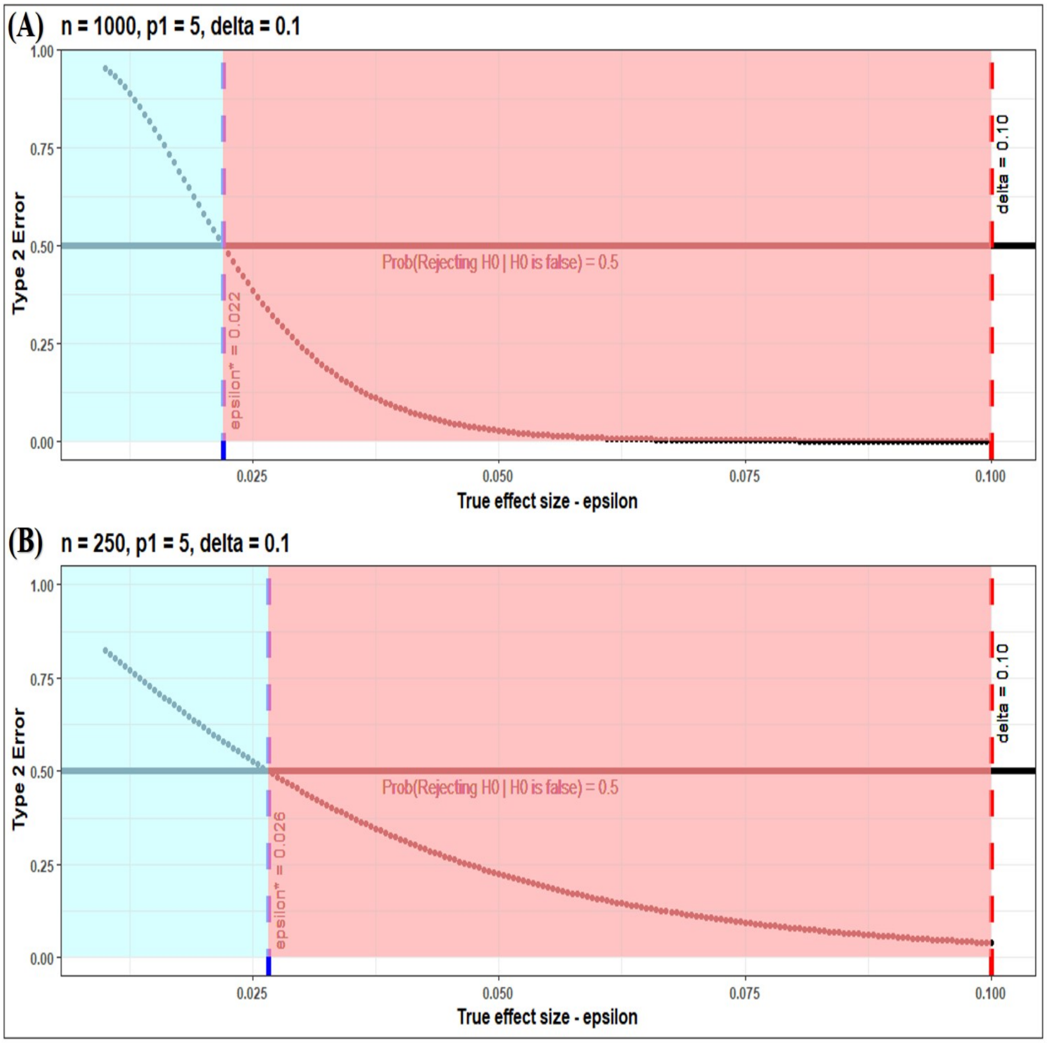

Figure 1: Null and alternative neighborhoods induced through neutral effect size and corresponding type 2 error function for sample sizes $n = 250$ and $n = 1000$

Table 1: type 1 and type 2 error rates for error calibrated cutoff

| $\boldsymbol{R^2}$ (baseline model) | $p_1$ | $f^2$ | $n$ | $\text{T}_{\text{ECC}}$ | $\alpha(\text{T})$ | $\beta(\text{T})$ |
|---|---|---|---|---|---|---|
| 0.2 | 1 | 0.02 | 200 | 1.508 | 0.267 | 0.200 |
| 0.2 | 1 | 0.04 | 200 | 2.560 | 0.133 | 0.067 |
| 0.2 | 1 | 0.06 | 200 | 3.588 | 0.067 | 0.033 |
| 0.2 | 1 | 0.02 | 350 | 2.299 | 0.312 | 0.245 |
| 0.2 | 1 | 0.04 | 350 | 4.097 | 0.187 | 0.167 |
| 0.2 | 1 | 0.06 | 350 | 5.870 | 0.091 | 0.091 |
| 0.2 | 1 | 0.02 | 500 | 3.075 | 0.089 | 0.089 |
| 0.2 | 1 | 0.04 | 500 | 5.618 | 0.044 | 0.052 |
| 0.2 | 1 | 0.06 | 500 | 8.137 | 0.001 | 0.002 |
| 0.2 | 3 | 0.02 | 200 | 4.001 | 0.267 | 0.486 |
| 0.2 | 3 | 0.04 | 200 | 5.058 | 0.171 | 0.199 |
| 0.2 | 3 | 0.06 | 200 | 6.017 | 0.086 | 0.057 |
| 0.2 | 3 | 0.02 | 350 | 4.806 | 0.338 | 0.242 |
| 0.2 | 3 | 0.04 | 350 | 6.475 | 0.152 | 0.191 |
| 0.2 | 3 | 0.06 | 350 | 8.005 | 0.031 | 0.000 |
| 0.2 | 3 | 0.02 | 500 | 5.546 | 0.111 | 0.101 |
| 0.2 | 3 | 0.04 | 500 | 7.792 | 0.074 | 0.059 |
| 0.2 | 3 | 0.06 | 500 | 9.872 | 0.014 | 0.000 |
